# The specificity of Japanese PCR assays for SARS-CoV-2 exceeds 99.7%

**DOI:** 10.1101/2020.05.15.20103515

**Authors:** Akihiro Tsukadaira, Kenichi Nishie

## Abstract

The Japanese government arranged a total of three charter flights to evacuate Japanese residents out of Wuhan Jan 29, 30, and 31, 2020. The project to rescue Japanese residents can be recognized as a randomized sampling study for the COVID-19 epidemic in Wuhan at that time. Jan 23, Jonathan MR et al stated in the medRxiv, in 14 days’ time (Feb 4, 2020), the number of infected people in Wuhan is estimated to be greater than 190 thousand (prediction interval, 132,751 to 273,649). In our study, the estimation of the COVID-19 cases in Wuhan around Jan 29, 30 and 31, 2020 exceeds the lower prediction 132,751 in the prior probability: 0.0134. Our statistical analysis is consistent with Jonathan MR et al, if the specificity of SARS–CoV–2 PCR assays in Japan exceeds 99.7%.

## Article type

### Letter to the Editor

Since the outbreak of the new coronavirus infection (COVID-19) in Wuhan became serious in the beginning of 2020, the Japanese government arranged charter flights to evacuate Japanese residents out of Wuhan. A total of three rescue planes returned from Wuhan Jan 29, 30, and 31, 2020. All 566 individuals rescued from Wuhan underwent SARS–CoV–2 PCR examinations, 7 passengers tested positive on arrival and were placed under quarantine, while 5 more passengers tested positive during the 2 weeks of health observation (1) (1). The breakdown of all returnees are listed in Table 1..

**Table 1:**
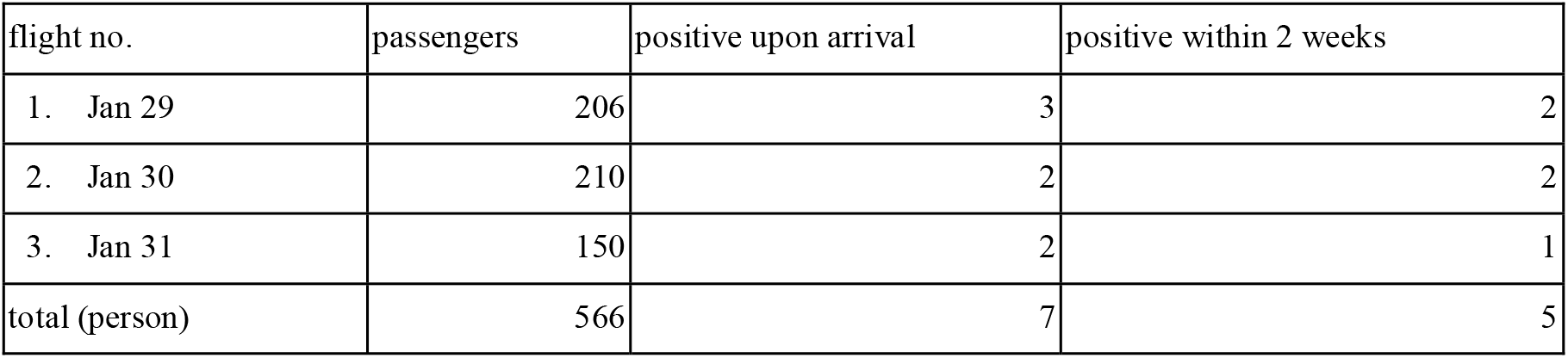
The breakdown of the 3 flights for Japanese residents evacuated from Wuhan on Jan 29, 30,and 31, 2020

| flight no. | passengers | positive upon arrival | positive within 2 weeks |
| --- | --- | --- | --- |
| 1. Jan 29 | 206 | 3 | 2 |
| 2. Jan 30 | 210 | 2 | 2 |
| 3. Jan 31 | 150 | 2 | 1 |
| total (person) | 566 | 7 | 5 |

In 2003 RT-PCR assays, from the World Health Organization (WHO) severe acute respiratory syndrome (SARS) network laboratories (at the University of Hong Kong and at the Bernhard-Nocht Institute in Hamburg, Germany) exhibited diagnostic sensitivities of 61% and 68% (nasopharyngeal aspirate specimens), 65% and 72% (throat swab specimens), respectively (3). The National Institute of Infectious Diseases (NIID) in Japan states that the PCR assays for SARS-CoV-2 is highly specific, but the exact sensitivity and specificity are still unknown.

The project to rescue Japanese residents can be recognized as a randomized sampling study for the COVID-19 epidemic in Wuhan at that time. The prior probability of statistics in Wuhan is unknown, however the following formula can be used for the back calculation.Table 2 shows statistical probability formulas assuming the sensitivity of SARS–CoV–2 PCR assays for nasopharyngeal and throat swab is 70%.

**Table 2:** Formula of prior probability when the sensitivity is 0.7 and the unknown specificity is a.Sensitivity: 0.7 Specificity: a 0.7х + (1–a) (566–х)= 7 The true patient count (positives and false negatives): х Prior probability: х/566

|  | patient | non-patient | total (person) |
| --- | --- | --- | --- |
| PCR (+) | $0.7x$ | $(1-a)(566-x)$ | 7 |
| PCR (-) | $0.3x$ | $a(566-x)$ | 559 |
| Total (person) | x | $566-x$ | 566 |

As of Jan 31, 2020, 192 deaths were reported, and 3215 positive COVID-19 cases were confirmed COVID-19 by examination in Wuhan (4) (5). If the specificity is 99.00 to 99.99%, a projected estimate of COVID-19 cases in Wuhan, a city with a population of 11 million, is shown on Table 3.. Jan 23, 2020, Jonathan MR et al stated in the medRxiv, in 14 days’ time (Feb 4, 2020), the number of infected people in Wuhan is estimated to be greater than 190 thousand (prediction interval, 132,751 to 273,649) (6).

**Table 3:** A projected estimate of COVID-19 cases in Wuhan to use as the prior probability when the sensitivity is 99.00 to 99.99%. The underlined numbers exceed the reported lower prediction interval.

| sensitivity (%) | specificity (%) | prior probability | patient in Wohan ( x 10,000 ) |
| --- | --- | --- | --- |
| 70 | 99.0 | 0.0034 | 3.7 |
| 70 | 99.1 | 0.0049 | 5.4 |
| 70 | 99.2 | 0.0063 | 6.9 |
| 70 | 99.3 | 0.0077 | 8.5 |
| 70 | 99.4 | 0.0092 | 10.1 |
| 70 | 99.5 | 0.0106 | 11.7 |
| 70 | 99.6 | 0.0120 | 13.2 |
| 70 | 99.7 | 0.0134 | <u>14.7</u> |
| 70 | 99.8 | 0.0149 | <u>16.4</u> |
| 70 | 99.9 | 0.0163 | <u>17.9</u> |
| 70 | 99.99 | 0.0175 | <u>19.3</u> |

In our study, the estimation of the COVID-19 cases in Wuhan around Jan 29, 30 and 31, 2020 exceeds the lower prediction 132,751 in the prior probability: 0.0134. Our statistical analysis is consistent with Jonathan MR et al, if the specificity of SARS-CoV-2 PCR assays in Japan exceeds 99.7%. If the specificity in Japan was 99.99%, it would almost match the number of greater than 190 thousand, Jonathan MR et al said.

## Data Availability

1. National Institute of Infectious Diseases in Japan Open Data [Internet]. 2020 [cited 2020 Apr 7]. Available from: https://www.niid.go.jp/niid/ja/diseases/ka/corona-virus/2019-ncov/2488-idsc/iasr-news/9525-483p01.html
2. National Institute of Infectious Diseases in Japan Open Data [Internet]. 2020 [cited 2020 Apr 10]. Available from: https://www.niid.go.jp/niid/ja/diseases/ka/corona-virus/2019-ncov/2488-idsc/iasr-news/9557-483p04.html

## Acknowledgments

A.T. and K.N. received no funding

## Authors’ contributions

All authors conceived the study and participated in the study design. A.T.collected and analysed the data. A.T. and K.N. drafted the manuscript and revised the the earlier versions of the manuscript. All authors edited the manuscript and approved the final version.

## Conflict of interest

The authors declare no conflicts of interest.

## Notes

### Competing Interest Statement

The authors have declared no competing interest.

### Clinical Trial

No trial ID for retrospective study

